# Biomarker changes preceding symptom onset in genetic prion disease

**DOI:** 10.1101/2023.12.18.23300042

**Authors:** Sonia M Vallabh, Meredith A Mortberg, Shona W. Allen, Ashley C Kupferschmid, Pia Kivisäkk, Bruno L Hammerschlag, Anna Bolling, Bianca A. Trombetta, Kelli Devitte-McKee, Abaigeal M. Ford, Lauren Sather, Griffin Duffy, Ashley Rivera, Jessica Gerber, Alison J McManus, Eric Vallabh Minikel, Steven E Arnold

## Abstract

**Importance:** Genetic prion disease is a universally fatal and rapidly progressive neurodegenerative disease for which genetically targeted therapies are currently under development. Preclinical proofs of concept indicate that treatment before symptoms will offer outsize benefit. Though early treatment paradigms will be informed by the longitudinal biomarker trajectory of mutation carriers, to date limited cases have been molecularly tracked from the presymptomatic phase through symptomatic onset.

**Objective:** To longitudinally characterize disease-relevant cerebrospinal fluid (CSF) and plasma biomarkers in individuals at risk for genetic prion disease up to disease conversion, alongside non-converters and healthy controls.

**Design, setting, and participants:** This single-center longitudinal cohort study has followed 41 *PRNP* mutation carriers and 21 controls for up to 6 years. Participants spanned a range of known pathogenic *PRNP* variants; all subjects were asymptomatic at first visit and returned roughly annually. Four at-risk individuals experienced prion disease onset during the study.

**Main outcomes and measures:** RT-QuIC prion seeding activity, prion protein (PrP), neurofilament light chain (NfL) total tau (t-tau), and beta synuclein were measured in CSF. Glial fibrillary acidic protein (GFAP) and NfL were measured in plasma.

**Results:** We observed RT-QuIC seeding activity in the CSF of three E200K carriers prior to symptom onset and death, while the CSF of one P102L carrier remained RT-QuIC negative through symptom conversion. The prodromal window of RT-QuIC positivity was one year long in an E200K individual homozygous (V/V) at PRNP codon 129 and was longer than two years in two codon 129 heterozygotes (M/V). Other neurodegenerative and neuroinflammatory markers gave less consistent signal prior to symptom onset, whether analyzed relative to age or individual baseline. CSF PrP was longitudinally stable (mean CV 10%) across all individuals over up to 6 years, including at RT-QuIC positive timepoints.

**Conclusion and relevance:** In this study, we demonstrate that at least for the E200K mutation, CSF prion seeding activity may represent the earliest detectable prodromal sign, and that its prognostic value may be modified by codon 129 genotype. Neuronal damage and neuroinflammation markers show limited sensitivity in the prodromal phase. CSF PrP levels remain stable even in the presence of RT-QuIC seeding activity.

**KEY POINTS:** *Question:* What biofluid-based molecular changes precede symptom onset in genetic prion disease? For any observed changes, how consistently and by how long do they precede onset?

*Findings:* In this longitudinal study, presymptomatic CSF RT-QuIC prion seeding activity was observed in three E200K carriers who subsequently developed and died of prion disease, with changes in prodromal timing noted based on *PRNP* codon 129 genotype. CSF PrP levels were longitudinally stable in all study participants up to 6 years, regardless of mutation status or presence of RT-QuIC seeding activity.

*Meaning:* These findings suggest that CSF RT-QuIC may offer a brief, genotype-dependent prodromal signal prior to symptom onset in carriers of the E200K mutation. They further indicate that across *PRNP* mutations, CSF PrP levels are sufficiently stable to serve as a drug activity biomarker for PrP-lowering therapeutics and will not be confounded by disease-related molecular changes that precede symptom onset.

## INTRODUCTION

Prion disease features striking biomarker signatures^1–4^, but limited data exist on pre-symptomatic changes^5–7^. Mirroring disease duration^8^, prodomal change in genetic prion disease appears brief, preceding symptoms by at most 1-4 years^6,7^. Prion “seeds” in CSF have been detected by real-time quaking induced conversion (RT-QuIC) in pre-symptomatic individuals^5,7^, but prognostic value remains unclear. Here, we report fluid biomarker trajectories associated with 4 disease onsets over 6 years in a longitudinal natural history of genetic prion disease mutation carriers.

## METHODS

### Study participants

This previously described^5^ cohort study includes asymptomatic individuals with pathogenic *PRNP* mutations; individuals at risk for same; and controls (Table 1; Figure S1). Individuals with contraindication to lumbar puncture were excluded. Each visit included CSF and plasma collection, a medical history and physical, and a battery of cognitive, psychiatric, and motor tests and inventories. Individuals were invited to complete a baseline visit, a short-term repeat 2-4 months later (pre-2020), and approximately yearly visits thereafter. Data presented here were collected July 2017 to February 2023 and include data previously reported^5,9^. All participants were cognitively normal and provided written informed consent. This study was approved by the Mass General Brigham Institutional Review Board (2017P000214). Assay validation utilized samples from MIND Tissue Bank (2015P000221).

**Table 1.** Demographic characteristics of the cohort. “Age” represents age last seen, follow-up is years from first visit to last visit, and both are represented by mean ± SD.

| mutation status | N | sex | age (y) | follow-up (y) | total visits | CSF samples | plasma samples | mutations |
| --- | --- | --- | --- | --- | --- | --- | --- | --- |
| carrier | 41 | 13M / 28F | 47.5±14.0 | 2.0±1.9 | 126 | 104 | 109 | 6 P102L<br>7 D178N<br>22 E200K<br>6 other |
| control | 21 | 6M / 15F | 46.1±13.3 | 1.4±1.5 | 57 | 51 | 51 | 21 none |

### Biomarker assays

Biomarker assays utilized were: RT-QuIC (IQ-CSF protocol)^10^, PrP ELISA^9^, Simoa (Quanterix) GFAP, and Ella (Bio-Techne) NfL, T-tau (Figure S3), and β-syn (Figure S4), see Supplementary Methods.

### Statistical analysis

Biomarker relationships with age and mutation status were assessed by log-linear regression; curve fits shown in figures are the separate best fits for mutation carriers and for controls, while P values are for the effect of carrier status in a combined model: lm(log(value) ∼ age + carrier). Our study does not disclose biomarker values or *PRNP* mutation status to participants, yet a combination of age and the number and spacing of visits completed could uniquely identify some individuals, presenting a self-identification risk. To mitigate this risk, for controls and non-converting carriers in data visualizations, ages were obfuscated by addition of a normally distributed random variable with mean of 0 and standard deviation of ±3 years, and visit spacing intervals were obfuscated by multiplication by a normally distributed random variable with mean 1 and standard deviation ±25%, capped at a maximum increase of +25% to avoid visually exaggerating the study’s duration. True ages and true visit intervals for all participants are used in all descriptive statistics and statistical models and true ages and true visit intervals are shown in plots for the individuals who converted to active disease. For details of RT-QuIC analysis see Supplementary Methods. P values <0.05 were considered nominally significant. Analyses were conducted in R 4.2.0. Source code, summary statistics for all participants, and individual biomarker values for converting participants are available at https://github.com/ericminikel/mgh_prnp_freeze2

## RESULTS

Of 41 carriers (Table 1), four converted to active disease (N=3 E200K, N=1 P102L). 6 RT-QuIC positives (Figure 1A) belonged to 3 E200K individuals who converted and died of prion disease. 2 *PRNP* codon 129 heterozygotes (M/V) were RT-QuIC positive at first sample (2.5 and 3.1 years before onset); prion titer in CSF did not appreciably rise thereafter (Figure 1B). One homozygote (V/V) became RT-QuIC positive on study and became symptomatic 1 year later.

**Figure 1.**
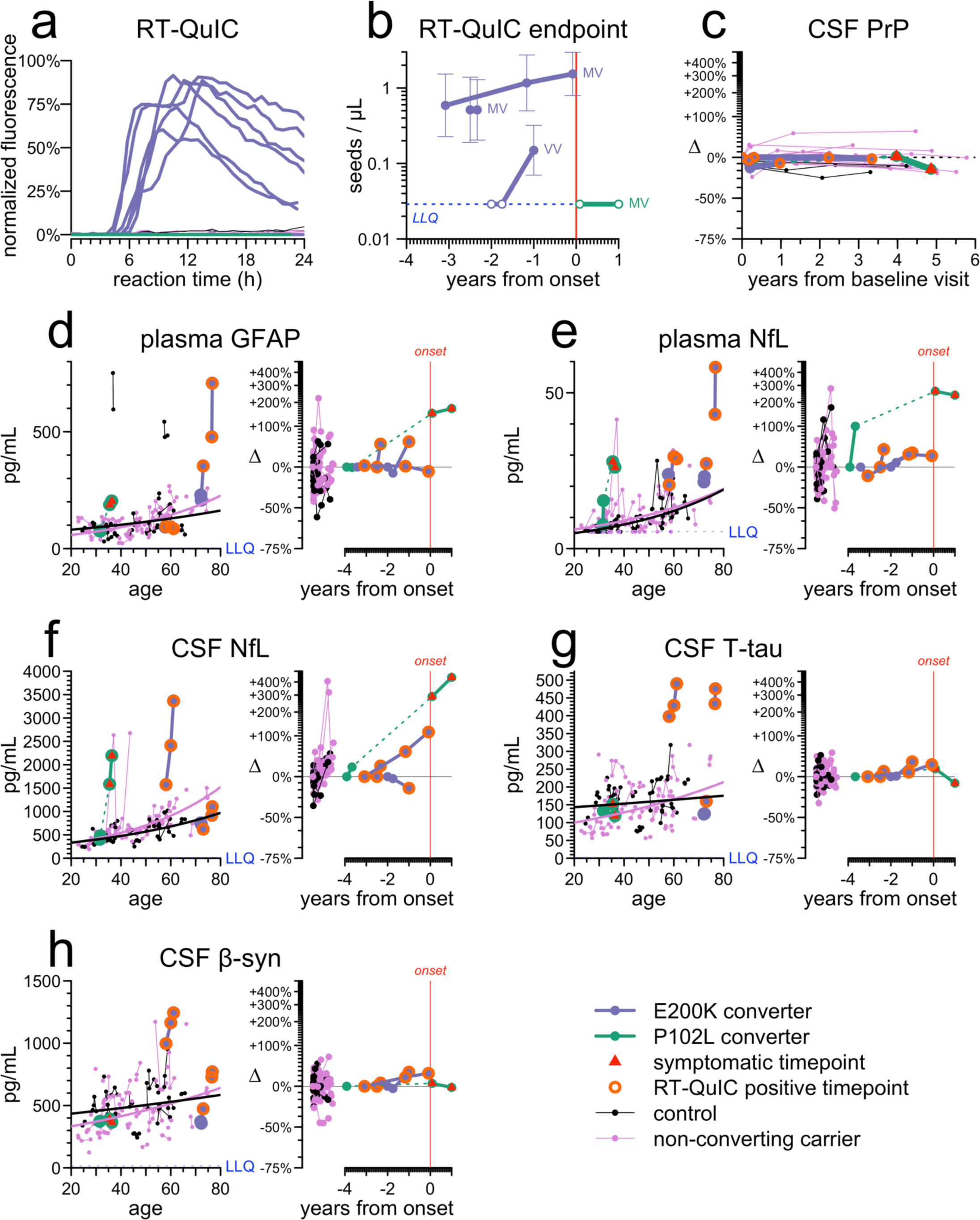
Fluid biomarker changes in the cohort. **A)** RT-QuIC kinetic curves, showing 6 positive CSF samples (each with 4/4 replicates positive) out of 149 tested (98 from carriers, 51 from controls). **B)** RT-QuIC endpoint titration of CSF, with codon 129 genotypes of converters indicated. **C)** CSF PrP concentrations represented as changes (Δ) relative to individual baseline, shown for the 4 converters plus all individuals (N=11) with at least 3 years between first and last available CSF sample. **D-H)** Biomarkers plasma GFAP (**D**), plasma NfL (**E**), CSF NfL (**F**), CSF T-tau (**G**), and CSF β-syn (**H**) are represented by two views each. Left: individual age vs. absolute concentration in pg/mL, with sequential samples from the same individual connected by thin lines, while thicker lines represent the separate log-linear best fit curves for controls and for non-converting carriers. Right: years from disease onset vs. change (Δ) relative to individual baseline in converters, with the same for controls and for non-converting carriers shown on a separate x-axis. Dashed lines connect timepoints before and after symptom onset.

CSF total PrP levels varied between individuals and were lower in mutation carriers (Figure S2) but were longitudinally stable in each individual out to 6 years (mean CV 10%) (Figure 1C), including samples taken after RT-QuIC positivity.

Plasma GFAP, a marker of reactive astrogliosis, was high relative to age in 2/4 converters, but change from individual baseline was unremarkable compared to controls and non-converters (Figure 1D). Plasma NfL appeared high and increased in all 4 converters, but not outside the range of non-converters and controls (Figure 1E). CSF NfL, CSF t-tau, and CSF beta-synuclein were each elevated in 2/4 converters and normal in 2/4 (Figure 1F-H); different converting individuals were high for different markers.

## DISCUSSION

Here we describe fluid biomarker profiles in a longitudinal cohort of genetic prion disease mutation carriers, including 4 individuals who converted to active disease. As before^5–7^, at any given time, cross-sectionally, most carriers of genetic prion disease mutations do not have any detectable molecular sign of disease. Our data support the hypothesis that CSF prion seeding activity as assayed by RT-QuIC may represent the first detectable change in E200K carriers. However, we did not detect seeding activity in the CSF of a P102L converter, consistent with RT-QuIC’s lower sensitivity for most non-E200K genetic subtypes^1,11^. Though our sample is small, our data suggest that PRNP codon 129 genotype may modify the duration of CSF RT-QuIC positivity before onset in E200K individuals; longer prodromal positivity in M/V heterozygotes would mirror their longer disease duration after onset^12^.

Soluble PrP in CSF is reduced in symptomatic prion disease patients, presumably as a result of a disease sink process^13–16^, and yet pharmacologic lowering of CSF PrP may be important as a drug activity biomarker for trials of PrP-lowering drugs, and has been proposed as a surrogate endpoint in prevention trials^17^. Our data suggest that CSF PrP does not begin to decline prior to symptom onset, even in the presence of RT-QuIC positivity, suggesting its use in asymptomatic individuals will not be confounded.

Neuronal damage and neuroinflammation markers rise with age and may vary between individuals. Neither when normalized to age nor to individual baseline did any of these markers consistently provide distinctive signal in all 4 of our converting individuals relative to non-converters and controls. Thus, while these markers may be useful as an adjunct, none is likely to provide the prognostic specificity of RT-QuIC. RT-QuIC, meanwhile, may offer just 1 year of advance signal in some E200K cases, and currently faces limited sensitivity to other subtypes. Assay improvement, biomarker discovery, and continued sample accrual will be vital to identifying additional prognostic markers, particularly for non-E200K subtypes. At any given time, most carriers appear non-prodromal, thus, in this rare disease, prodromal individuals are unlikely to be identified in sufficient numbers to power clinical trials. Instead, primary prevention trials with inclusion based on genotype and CSF PrP as primary endpoint may be necessary^17^, and would honor the outsize benefit of early treatment observed in animal models^18^. Treatment of prodromal individuals could feature as a supportive arm and/or randomization off-ramp for carriers who develop a prodromal signature during a trial.

### Limitations

Four symptom onsets is a small absolute number from which to draw conclusions. Reflecting study enrollment and overall mutation prevalence, our observed onsets are skewed towards E200K. Some annual visits were missed due to COVID-19. We did not collect emerging sample types such as nasal brushings^19^, urine^20^, or tears^21^. Additional pre-symptomatic natural history work across multiple sites^7,22,23^ will be required to build confidence in our observations.

## CONCLUSIONS

In E200K carriers, RT-QuIC seeding activity in CSF can precede symptom onset by 1-3 years, perhaps depending on *PRNP* codon 129 genotype. CSF and plasma markers of neurodegeneration and neuroinflammation do not unambiguously identify imminent converters. CSF PrP levels are longitudinally stable over time in all participants even following RT-QuIC positivity.

## AUTHOR CONTRIBUTIONS

Dr. Vallabh had full access to all of the data in the study and takes responsibility for the integrity of the data and the accuracy of the data analysis. Concept and design: SMV, SEA, EVM. Patient visits and sample accrual: SEA, AJM, SWA, ACK, AMF, AB, KDM, GD. Sample analysis: MAM, SWA, PKW, BLH, AB, BAT. Supervision: SMV, AR, JG, AJM, EVM, SEA. Drafting of the manuscript: SV. Critical review of the manuscript: all authors. Statistical analysis: EVM. Obtained funding: SEA, SMV, EVM.

## Data Availability

All data produced in the present work are contained in the manuscript, supplement, and the study's Github repository, available online at https://github.com/ericminikel/mgh_prnp_freeze2

## SUPPLEMENT

### Supplementary Methods

#### Genotyping

Whole blood was frozen hemolyzed and genomic DNA was extracted. All samples were genotyped by two orthogonal methods. DNA was submitted for targeted capture using a custom set of probes (Twist Biosciences) directed against ∼150 kb of genomic sequence in and surrounding *PRNP*, then enriched DNA was subjected to deep short-read sequencing (Illumina) at the Broad Institute’s Genomics Platform. Data were aligned to the hg38 reference genome and processed using Dragen 3.7.8 to yield multi-sample VCF files. In parallel, DNA also underwent a previously described^24^ protocol implemented by Genewiz, combining Sanger sequencing to detect SNPs and short indels with gel sizing to detect octapeptide repeat insertions (OPRI).

Briefly, the primers utilized are: Int5: 5′-TgCATgTTTTCACgATAgTAACgg-3′, DG2: 5′-gCAgTCATTATggCgAACCTTggCTg-3′, and 3′Sal: 5′-gTACTgAggATCCTCCTCATCCCACTATCAggAAgA-3′. The prodcut of the DG2/3′Sal reaction is subjected to Sanger sequencing; the product of the DG2/Int5 reaction is run on a 2% agarose gel (the wild-type product is 464 bp). Genotypes obtained by the two different methods were in agreement for all samples. Determination of haplotypes was accomplished by molecular phasing of codon 129 to pathogenic variants by paired-end Illumina sequencing reads using a custom Python 3 script run on Terra (Terra.bio); source code is available in the study’s online GitHub repository. Our study includes individuals who are at risk for inheriting a *PRNP* mutation but have not undergone predictive testing; genotypes were determined for research purposes only and were not disclosed to participants.

#### Sample processing

Blood was collected in purple top K+ EDTA tubes, inverted gently, and centrifuged at 1,500 *g* for 10 minutes to retrieve plasma, aliquoted, and frozen at -80°C. 20 mL of CSF was collected via gentle aspiration lumbar puncture using a 24G atraumatic Sprotte needle into 4x 5 mL syringes. Because PrP in CSF is highly sensitive to polypropylene adsorption, we followed the protocol described in Vallabh 2019 Figure S8, where 2 of the 4 collected 5 mL aliquots were ejected into tubes pre-loaded with the zwitterionic detergent CHAPS (3% wt/vol stock solution at 1% volume to yield a final 0.03% CHAPS concentration). All CSF were centrifuged at 2,000 *g* for 10 minutes to remove cells, and then aliquoted and frozen at -80°C. In instances where the LP yielded only a limited volume of CSF, CHAPS aliquots were prioritized. CHAPS aliquots were used for PrP and NfL quantification. Neat aliquots were used for RT-QuIC. For T-tau and beta-synuclein, neat aliquots were used where available, while CHAPS aliquots were used when these were the only available samples; an assessment of the effect of 0.03% CHAPS on these assays is provided in Figures S3-S4. To minimize bias, technicians processing samples and performing biomarker assays were blinded to genotype.

#### RT-QuIC

Real-time quaking-induced conversion (RT-QuIC) was performed according to the protocol of Orru et al 2015, widely referred to as the IQ-CSF protocol^10^. The substrate was recombinant N-terminally truncated Syrian hamster PrP (SHaPrP90-230) expressed in *E. coli* and produced in-house according to a published protocol^25,26^ and filtered by centrifugation at 3,214 *g* through a 100 kDa filter (PALL OD100C33). Final concentration in the reaction was 300 mM NaCl (Broad Institute SQM), 10 mM sodium phosphate pH 7.4 (Molecular Toxicology; Thermo C790B91), 1 mM EDTA (Broad Institute SQM), 10 µM thioflavin T (Sigma T3516-5G), 0.002% sodium dodecyl sulfate (SDS) (Invitrogen 15553-035), and 0.1 mg/mL recombinant PrP, all diluted into distilled water (InvitroGen UltraPure 10977-015). 80 µL of a 1.25x concentrated master mix was loaded into each well of a 96-well plate (Nunc; Thermo 265301) and then 20 µL of CSF was added. Plates were sealed with adhesive film (VWR 37000-548). The assay was run at 55°C for 24 hours on a BMG FLUOStar OPTIMA platereader with alternating cycles of 1 minute rest and 1 minute 800 rpm shaking, with thioflavin T fluorescence measurements obtained via bottom read at 45-minute intervals with 450 nm excitation and 480 nm emission. Fluorescence kinetic curves were normalized so that 0% represents the baseline fluorescence value at first reading and 100% represents the instrument’s maximum value of 65,000 fluorescence units. We committed to the pre-specified criteria of Orru et al^10^: a CSF sample was called positive if at least 50% of technical replicates (e.g. 2/4) yielded at least 10% normalized signal within 24 hours. In practice, when screening undiluted CSF, all our positive samples were positive in 4/4 replicates while all negatives were positive in 0/4 replicates. For endpoint titration, 3-fold serial dilutions were run by adding 20, 6.7, 2.2, or 0.7 µL of CSF and then 0, 13.3, 17.8, or 19.3 µL of distilled water (InvitroGen UltraPure 10977-015). Titers were determined by Spearman-Karber analysis^27^; the source code is available in this study’s online GitHub repository.

#### PrP ELISA

PrP enzyme-linked immunosorbent assay (ELISA) was performed according to an in-house protocol previously published and described in detail^9^. The assay uses antibodies EP1802Y (Abcam ab52604) for capture and 8H4 (Abcam ab61409), biotinylated in-house, for detection. The standard curve is recombinant full-length mouse PrP (MoPrP23-231) produced in house, plated at concentrations from 0.05 ng/mL to 5 ng/mL. CSF was run at a dilution factor of 80, at which the lower limit of quantification (LLOQ) is 4 ng/mL. For longitudinal analysis (Figure 1C, Table S4), each individual was normalized to their own baseline. For comparison across mutations (Figure S2, Table S3), all individuals were normalized to the mean value in mutation-negative subjects, which was 70.6 ng/mL.

#### GFAP

Plasma GFAP was quantified using Simoa (Quanterix) according to manufacturer instructions at a dilution factor of 4, yielding an LLOQ of 2.744 pg/mL. Samples were run in technical duplicate with a mean CV of 6.0%.

#### NfL

Plasma and CSF NfL were quantified using Ella by ProteinSimple (Bio-Techne) at a dilution factor of 2 yielding an LLOQ of 5.4 pg/mL. CSF aliquots containing CHAPS were used. For all Ella assays, samples were plated onto cartridges in singlicate; each sample is then run in technical triplicate with three glass nanoreactors (GNRs).

#### T-tau

CSF T-Tau was analyzed both by ELISA (Fujirebio) and by Ella (Bio-Techne). For Ella, samples were run at a dilution factor of 2 (except for N=6 samples run at a dilution factor of 3 due to limited volume), with an LLOQ of 1.68 pg/mL. For ELISA, samples were run at a dilution factor of 4, with an LLOQ of 39.5 pg/mL. Ella results are reported in Figure 1G, while a comparison of the two assays is given in Figure S3.

#### Beta-synuclein

CSF beta-synuclein was analyzed by Ella (Bio-Techne) at a dilution factor of 2 for CSF (LLOQ: 15.9 pg/mL) and either 4 or 8 for plasma depending on available sample volume (LLOQ: 31.8 pg/mL or 63.7 pg/mL respectively). As shown in Figure S4, all plasma samples from study participants were at LLOQ.

## Supplementary Figures

**Figure S1.**
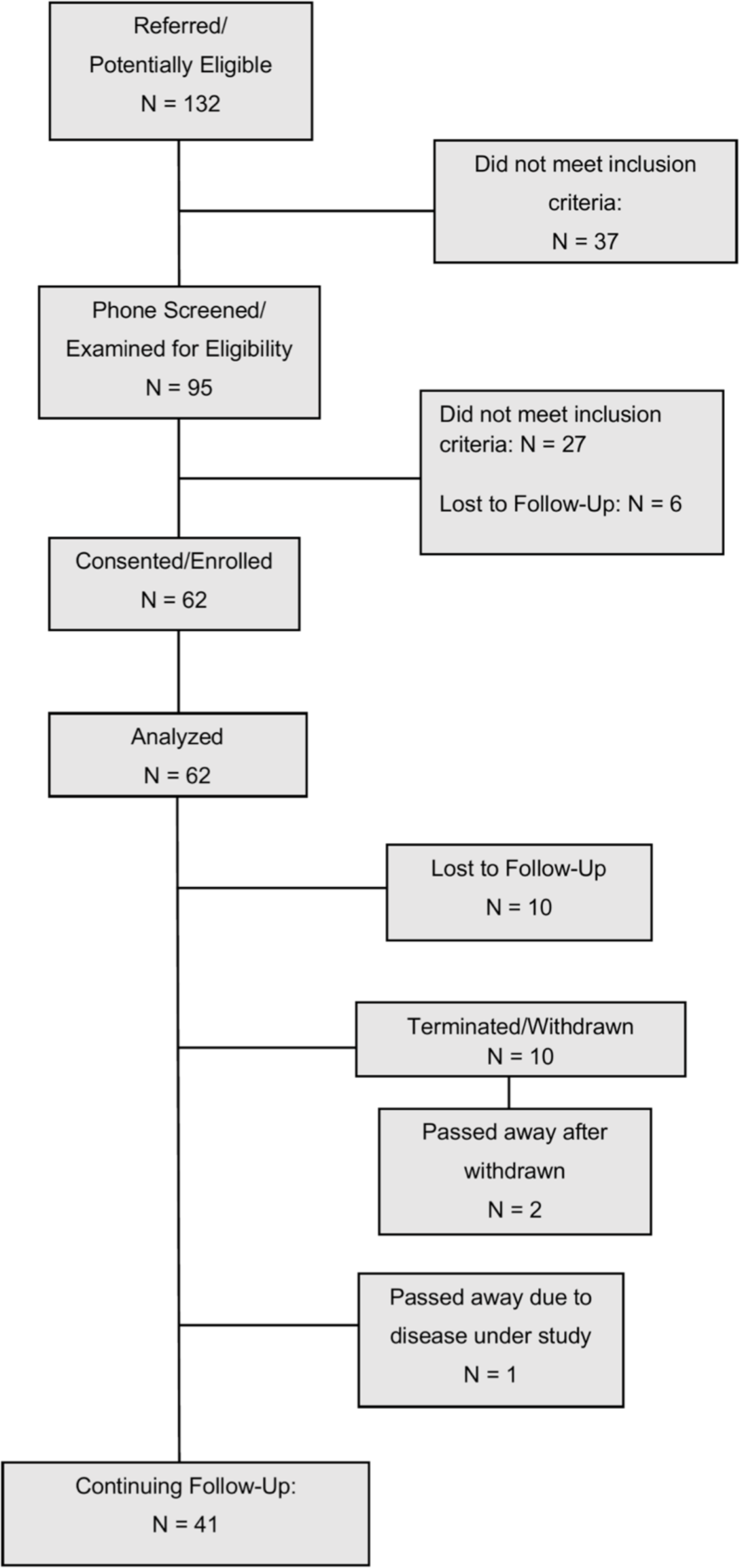
Flow chart of participant recruitment. At launch in July 2017, the study was open to known mutation carriers, those at risk, and known controls. From November 2021 new enrollment restricted to only known carriers, but already-enrolled individuals were invited to continue to participate regardless.

**Figure S2.**
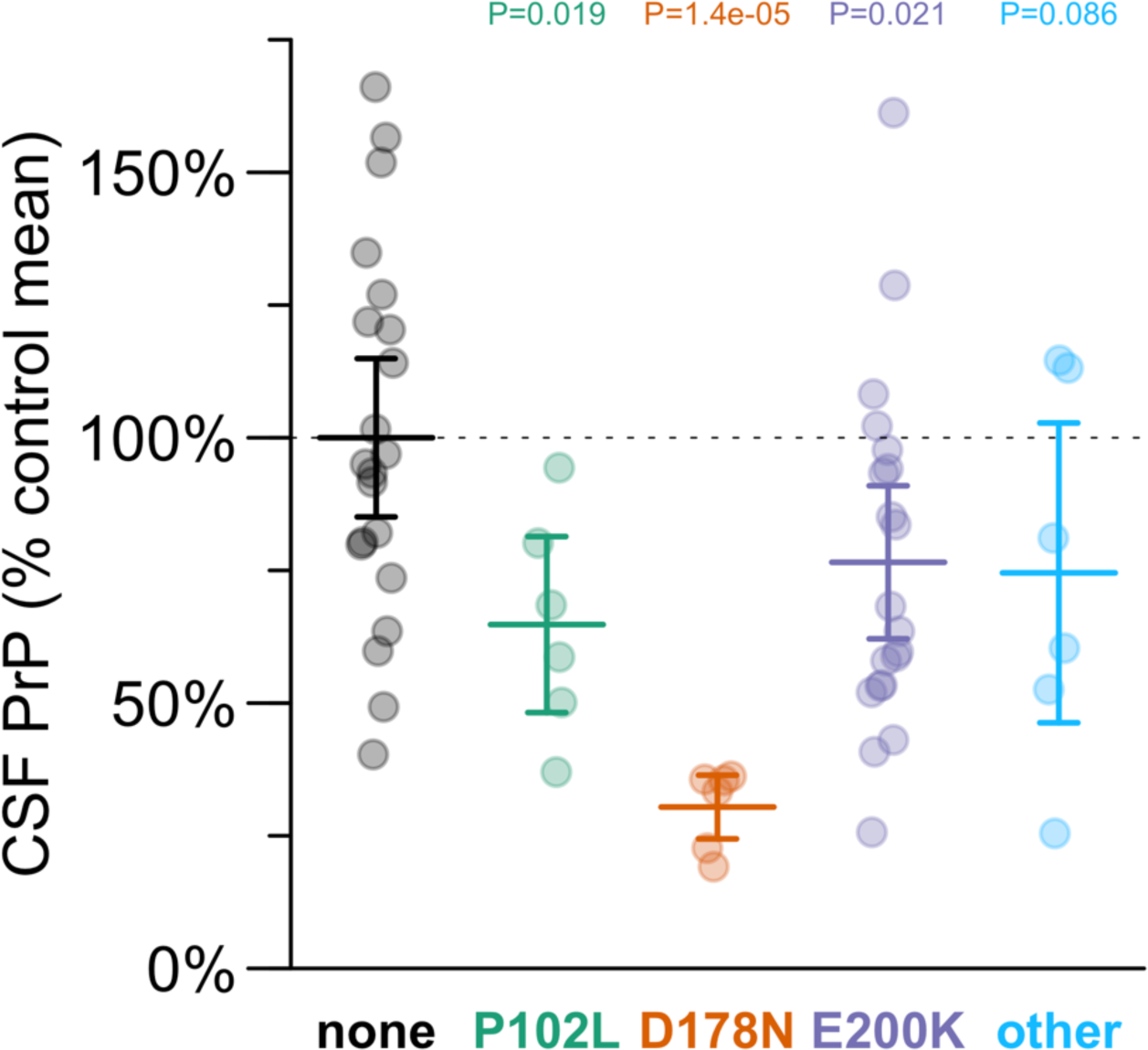
CSF PrP concentration by PRNP mutation. Each point represents the mean of all available CSF samples for one study participant. Data are normalized to the mean of the mutation-negative controls (“none”). P values are for differences from the control group in a linear model (lm in R, equivalent to Type I ANOVA).

**Figure S3.**
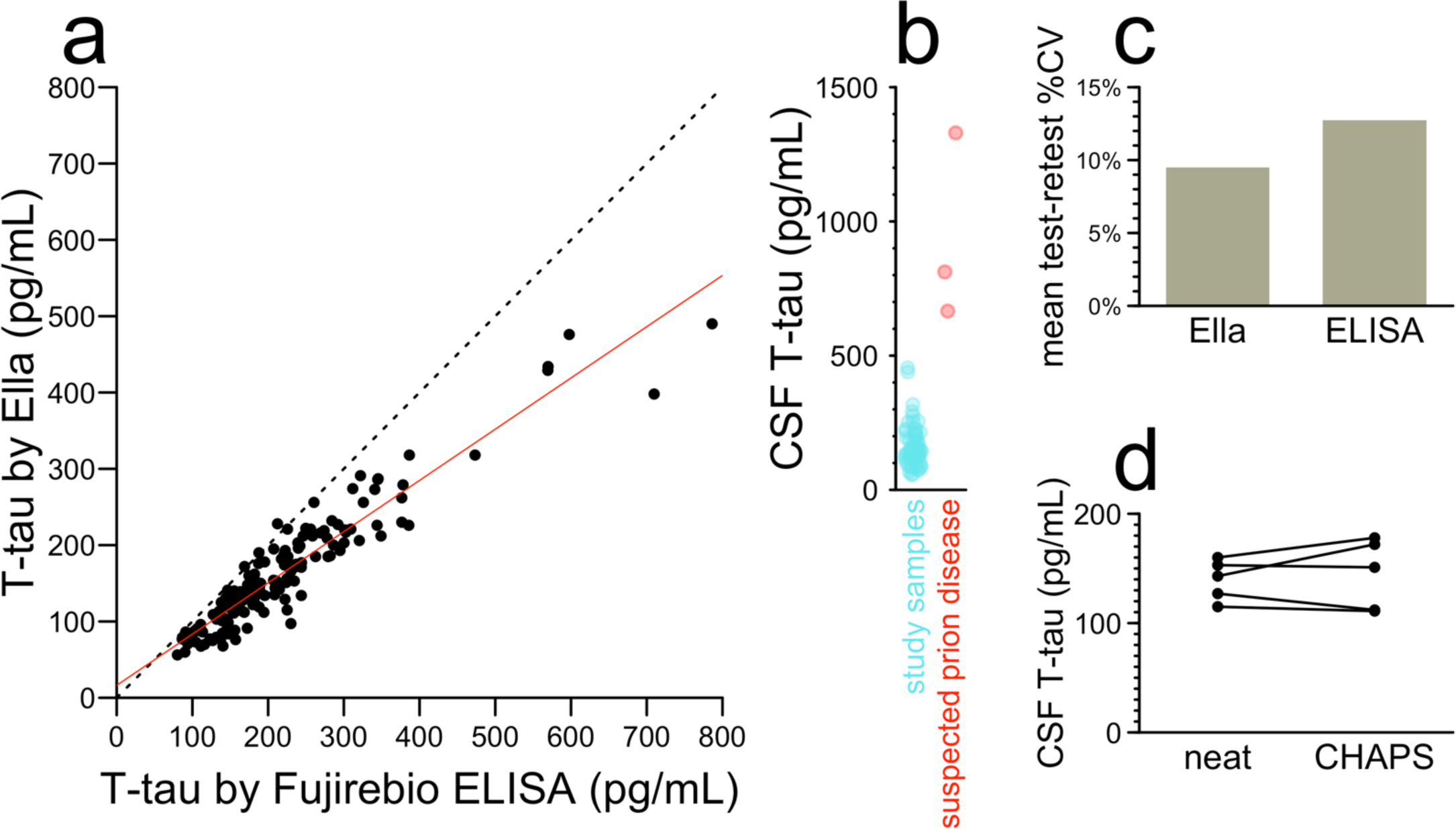
Quality control analyses on the Ella T-tau assay. **A)** Comparison of CSF T-tau concentrations in pg/mL for N=151 CSF samples determined by Fujirebio ELISA (x axis) versus Ella (y axis). The red line shows the best fit linear regression which is ella = 16.3 pg/mL + 67% × elisa. The Pearson’s correlation is r = 0.94, P = 5.3e-70. **B)** Comparison of mean CSF T-tau values per individual by Ella in study participants vs. 3 symptomatic patients with suspected prion disease. **C)** Mean test-retest CV for longitudinal LPs from the same individual: Ella 9.5%, ELISA 12.7%. **D)** Paired analysis of N=5 CSF samples analyzed by T-tau Ella both with and without the addition of 0.03% CHAPS. Mean value with CHAPS is 3% higher, P = 0.55 by paired T test.

**Figure S4.**
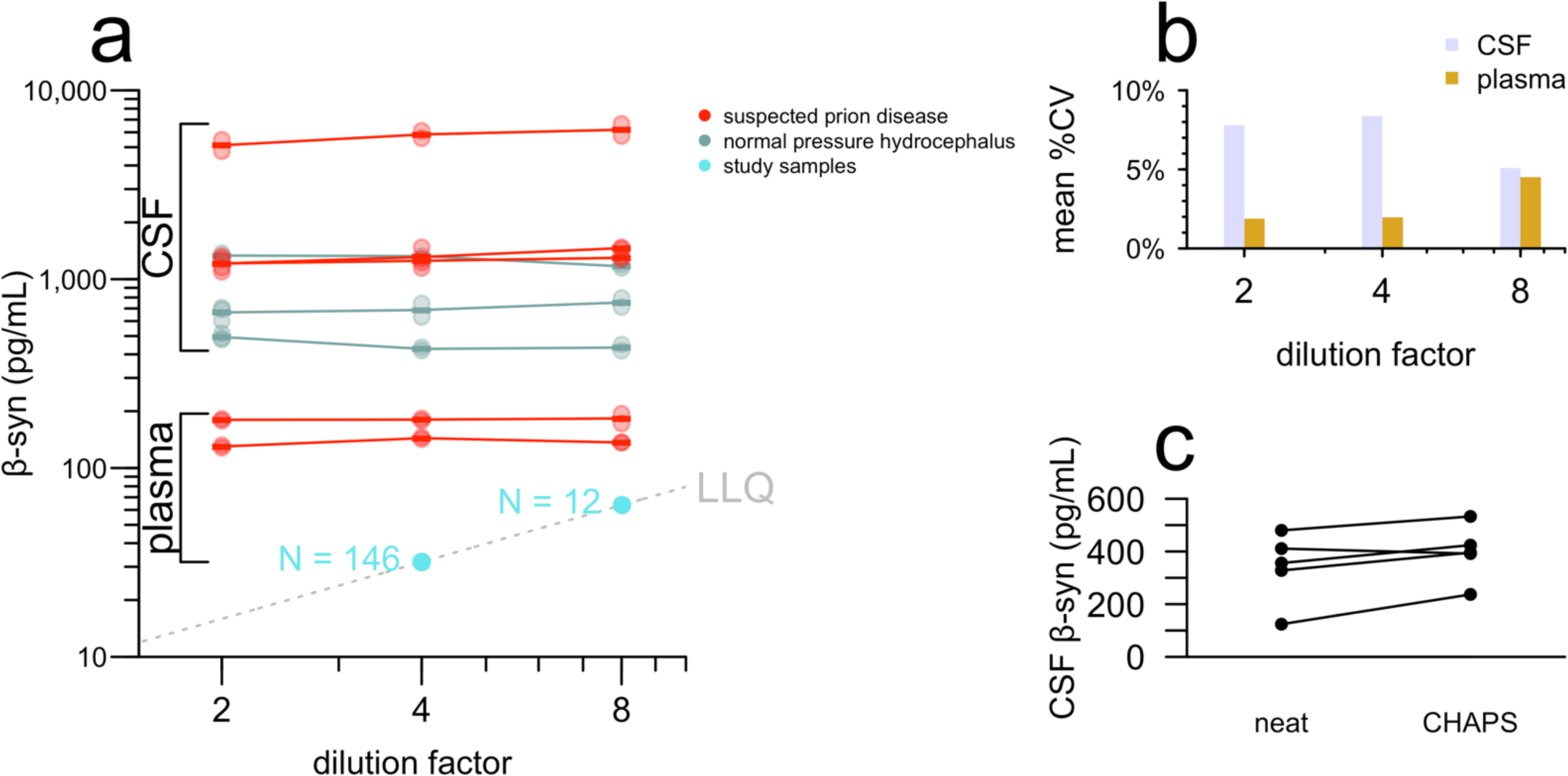
Quality control analyses on the Ella beta-synuclein assay. **A)** Parallelism (also called dilution linearity) tested on 6 CSF samples (3 suspected prion disease and 3 normal pressure hydrocephalus) and 2 plasma samples (2 suspected prion disease). Suspected prion disease patients are symptomatic individuals seen clinically at Massachusetts General Hospital outside of our study. All plasma samples from participants in our study were at the lower limit of quantification (LLQ). **B)** Mean coefficient of variation among 2 technical replicates per sample for the samples shown in (A). Note that this refers to plating the same sample twice, in separate wells, on the Ella cartridge; the measurement in each well is in turn the average of 3 replicate measurements. **C)** Comparison of 5 CSF samples from study participants analyzed both with and without the addition of 0.03% CHAPS, a detergent shown to reduce loss of PrP to plastic. Mean 27% higher reading in CHAPS samples, P = 0.057, paired T-test.

## Supplementary Tables

Supplementary tables are also available in the attached Excel spreadsheet and as tab-separated text files in the study’s online GitHub repository.

**Table S1.** All biomarker values from all visits by individuals who developed active disease. Genotype shows the pathogenic variant and codon 129. The 129MV individuals in this table are all cis-129M, trans-129V. Months from onset is negative for visits prior to symptom onset and positive for visits after symptom onset. Blank cells indicate assays not done because samples not collected (unsuccessful LP or virtual visit) or due to limited sample volume. MoCA^28^ and MRC Scale^29^ have been described elsewhere.

| Individual | Genotype | Age of onset bin | Visit number | Months from onset | CSF RT-QuIC | CSF RT-QuIC replicates | CSF T-tau (pg/mL) | CSF NfL (pg/mL) | CSF β-syn (pg/mL) | CSF PrP (ng/mL) | Plasma NfL (pg/mL) | Plasma GFAP (pg/mL) | MoCA score | MRC Scale |
| --- | --- | --- | --- | --- | --- | --- | --- | --- | --- | --- | --- | --- | --- | --- |
| A | E200K MV | 75-79 | 1 | -30 | + | 4/4 | 434 | 918 | 728 | 73.0 | 43.1 | 478.0 | 27 | 20 |
|  |  |  | 2 | -28 | + | 4/4 | 476 | 1103 | 771 | 69.8 | 58.2 | 707.7 | 25 | 20 |
| B | P102L MV | 35-39 | 1 | -47 |  |  |  | 406 | 373 | 37.5 | 7.7 | 75.4 | 26 | 20 |
|  |  |  | 2 | -44 |  |  | 133 | 478 |  | 33.9 | 15.4 | 74.8 | 27 | 20 |
|  |  |  | 3 | 1 | - | 0/4 | 152 | 1584 | 392 | 38.1 | 27.9 | 187.9 | 27 | 20 |
|  |  |  | 4 | 12 | - | 0/4 | 119 | 2194 | 366 | 30.6 | 26.1 | 204.0 | 28 | 19 |
| C | E200K VV | 65-69 | 1 | -24 | - | 0/4 | 124 | 757 | 372 | 32.8 | 21.4 | 229.6 | 29 | 20 |
|  |  |  | 2 | -21 | - | 0/4 | 125 | 732 | 358 | 27.6 | 23.1 | 207.2 | 28 | 20 |
|  |  |  | 3 | -12 | + | 4/4 | 160 | 623 | 474 | 29.9 | 27.3 | 353.7 | 24 | 20 |
| D | E200K MV | 60-64 | 1 | -41 |  |  |  |  |  |  | 23.8 | 91.8 | 26 | 20 |
|  |  |  | 2 | -37 | + | 4/4 | 398 | 1575 | 997 | 66.4 | 20.5 | 94.1 | 26 | 20 |
|  |  |  | 3 | -16 |  |  |  |  |  |  |  |  | 26 | 20 |
|  |  |  | 4 | -14 | + | 4/4 | 429 | 2413 | 1164 | 66.2 | 29.5 | 93.1 | 27 | 20 |
|  |  |  | 5 | -1 | + | 4/4 | 490 | 3365 | 1244 | 64.8 | 28.8 | 85.3 | 29 | 20 |

**Table S2.** Means, standard deviations, and ranges of biomarker values from all visits by participants who did not develop active disease, by mutation status. These summary statistics exclude all visits from the 4 participants who converted to active disease. In each cell, the top row shows mean±SD, while the bottom row shows range (min-max). CSF RT-QuIC positive shows the number of CSF samples that yielded an overall positive call. Each RT-QuIC reaction was run in quadruplicate; in this study, every positive sample was positive in all 4/4 replicates, while every negative sample was positive in 0/4 replicates.

| Group | N visits | CSF RT-QuIC positive | CSF T-tau (pg/mL) | CSF NfL (pg/mL) | CSF β-syn (pg/mL) | CSF PrP (ng/mL) | Plasma NfL (pg/mL) | Plasma GFAP (pg/mL) | MoCA | MRC |
| --- | --- | --- | --- | --- | --- | --- | --- | --- | --- | --- |
| Mutation-negative control | 57 | 0/51 | 164±49<br>(75-318) | 573±208<br>(242-1018) | 516±144<br>(243-952) | 71±24<br>(27-120) | 9.7±4.6<br>(5.4-28.2) | 145.9±150.1<br>(30.6-751.1) | 28±3<br>(14-30) | 20±0<br>(17-20) |
| Non-converting mutation carrier | 112 | 0/88 | 149±65<br>(56-318) | 685±404<br>(192-2677) | 472±201<br>(124-1171) | 45±24<br>(10-126) | 11.1±6.6<br>(5.4-41.4) | 112.8±53.3<br>(22.3-267.5) | 28±2<br>(22-30) | 20±0<br>(19-20) |

**Table S3.** Mean CSF PrP concentration (ng/mL) by mutation. These are the numeric values for the data shown in Figure S2. Results were grouped first by individual to determine mean CSF PrP concentration across longitudinal CSF samples, then grouped by mutation to determine mean and SD across individuals. N is the number of individuals in each group.

| Mutation | N | Mean | SD | Normalized mean | Linear regression P value |
| --- | --- | --- | --- | --- | --- |
| none | 21 | 69.9 | 24.4 | 100.0% | — |
| P102L | 6 | 45.3 | 14.5 | 64.8% | 1.92e-02 |
| D178N | 6 | 21.3 | 5.2 | 30.4% | 1.42e-05 |
| E200K | 20 | 53.5 | 23.0 | 76.5% | 2.06e-02 |
| other | 6 | 52.1 | 24.7 | 74.5% | 8.64e-02 |

**Table S4.**
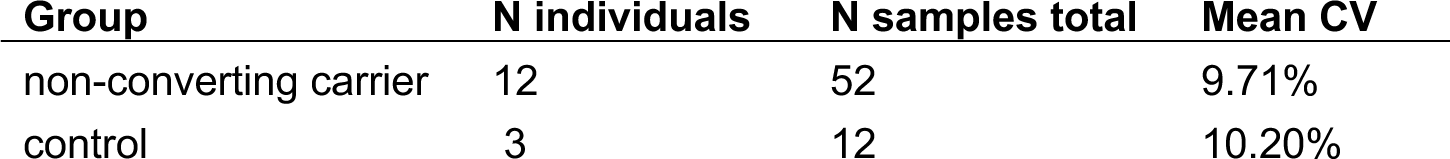
Long-term test-retest reliability of CSF PrP. Summary of data from Figure 1C. Test-retest mean CV summarized for all non-converting study participants with ≥3 years of longitudinal CSF data.

**Table S5.** Descriptive statistics and log-linear model fits on CSF and plasma biomarkers.

| Biomarker | N individuals | N samples | Carrier samples | Control samples | Test-retest mean CV without converters | Overall model<br>lm(log(value) ~ age + carrier, data = all) |  |  |  |  |
| --- | --- | --- | --- | --- | --- | --- | --- | --- | --- | --- |
|  |  |  |  |  |  | Y intercept | Annual increase | Annual increase P value | Carrier difference | Carrier difference P value |
| CSF NfL | 60 | 155 | 104 | 51 | 16.3% | 183.69 | 2.3% | 8.1e-20 | 0.0028 | 0.0028 |
| CSF T-tau | 60 | 151 | 100 | 51 | 9.6% | 101.29 | 0.9% | 2.8e-04 | 0.0734 | 0.0734 |
| CSF β-syn | 60 | 150 | 99 | 51 | 10.8% | 329.05 | 0.9% | 3.8e-04 | 0.0819 | 0.0819 |
| plasma NfL | 62 | 160 | 109 | 51 | 19.6% | 3.51 | 2.0% | 9.3e-14 | 0.0558 | 0.0558 |
| plasma GFAP | 61 | 158 | 107 | 51 | 18.4% | 45.78 | 1.9% | 6.3e-08 | 0.6014 | 0.6014 |

| Biomarker | Model of non-converting carriers only<br>lm(log(value) ~ age, data = NCcarriers) |  |  | Model of controls only<br>lm(log(value) ~ age, data = controls) |  |  |
| --- | --- | --- | --- | --- | --- | --- |
|  | Y intercept | Annual increase | Annual increase P value | Y intercept | Annual increase | Annual increase P value |
| CSF NfL | 193.9 | 2.6% | 2.7e-15 | 233.9 | 1.8% | 5.4e-06 |
| CSF T-tau | 77.5 | 1.3% | 2.8e-04 | 133.9 | 0.3% | 0.35 |
| CSF β-syn | 267.5 | 1.1% | 0.0014 | 393.6 | 0.5% | 0.13 |
| plasma NfL | 4.2 | 1.9% | 1.9e-07 | 3.2 | 2.3% | 1.0e-09 |
| plasma GFAP | 37.1 | 2.3% | 6.2e-10 | 64.1 | 1.2% | 0.11 |

## STROBE checklist

STROBE Statement—Checklist of items that should be included in reports of ***cohort studies***

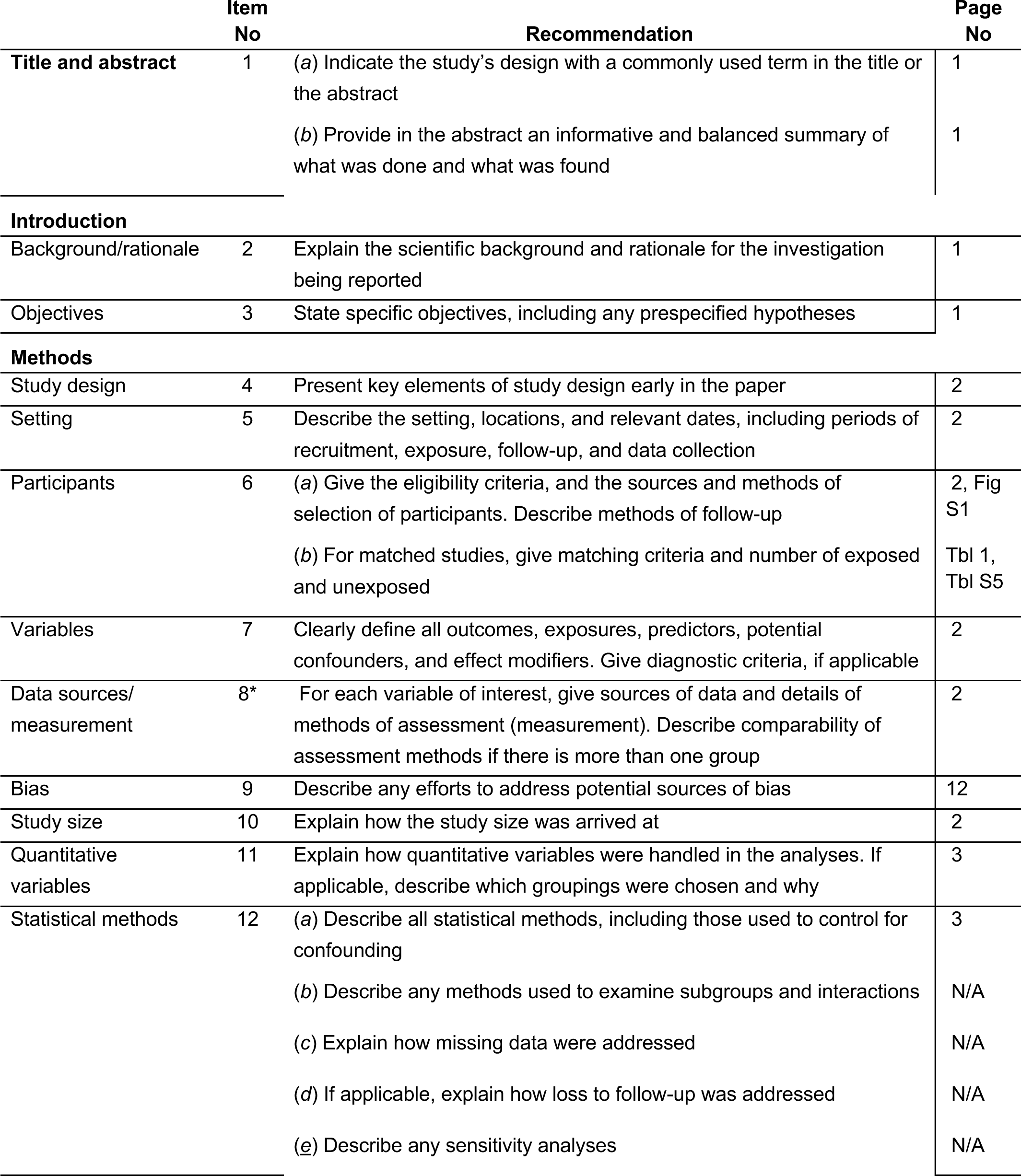
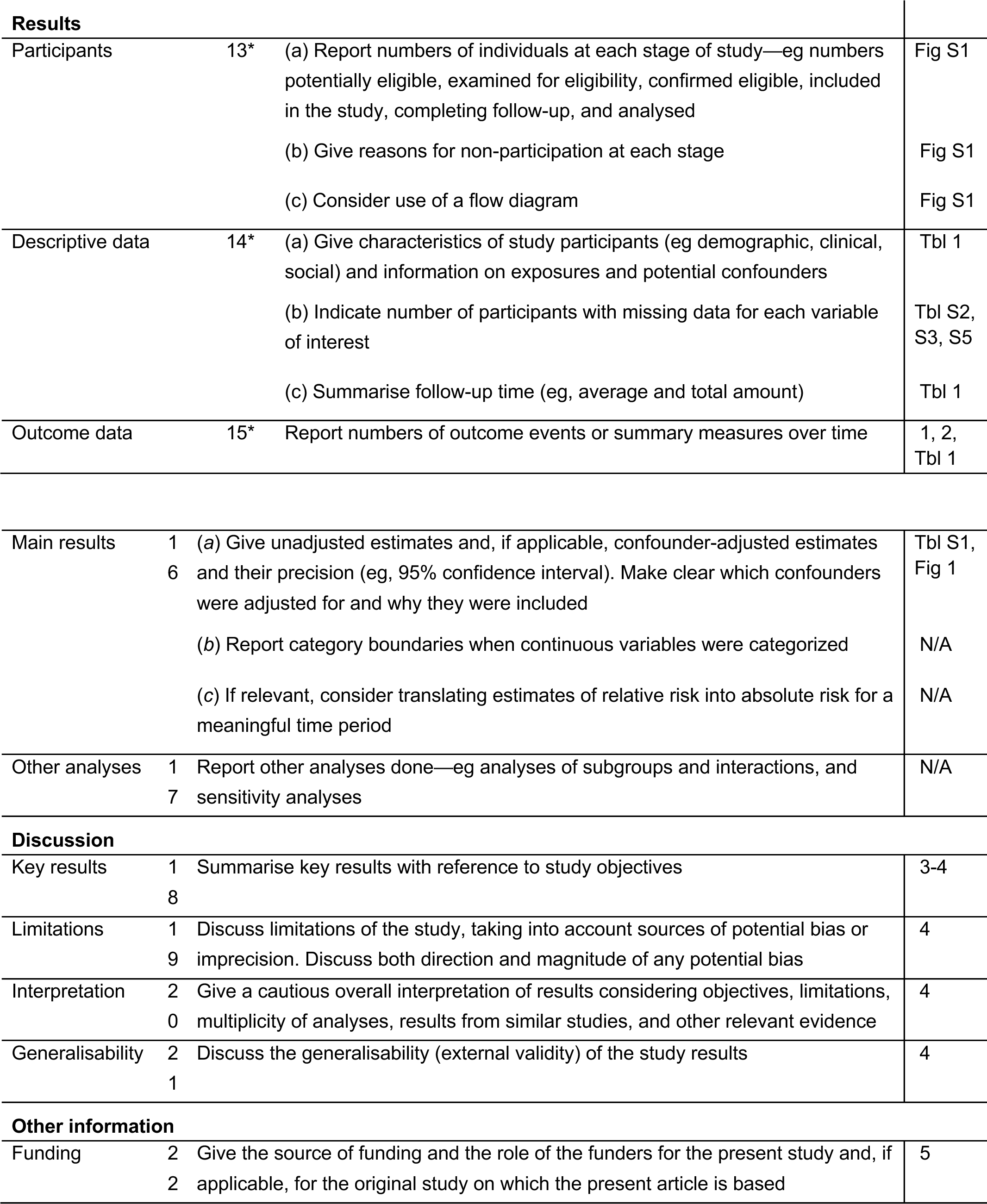

